# Mobility traces and spreading of COVID-19

**DOI:** 10.1101/2020.03.27.20045302

**Authors:** Sebastian Alexander Müller, Michael Balmer, Andreas Neumann, Kai Nagel

**Affiliations:** Transport Systems Planning and Transport Telematics, TU Berlin, Germany; Senozon AG, Switzerland; Senozon Deutschland GmbH, Germany

## Abstract

We use human mobility models, for which we are experts, and attach a virus infection dynamics to it, for which we are not experts but have taken it from the literature, including recent publications. This results in a virus spreading dynamics model. The results should be verified, but because of the current time pressure, we publish them in their current state. Recommendations for improvement are welcome. We come to the following conclusions:

1. **Complete lockdown works**. About 10 days after lockdown, the infection dynamics dies down. This assumes that lockdown is complete, which can be guaranteed in the simulation, but not in reality. Still, it gives strong support to the argument that it is never too late for complete lockdown.
2. As a rule of thumb, we would **suggest complete lockdown no later than once 10% of hospital capacities available for COVID-19 are in use**, and possibly much earlier. This is based on the following insights:
  a. Even after lockdown, the infection dynamics continues at home, leading to another tripling of the cases before the dynamics is slowed.
  b. There will be many critical cases coming from people who were infected before lockdown. Because of the exponential growth dynamics, their number will be large.
  c. Researchers with more detailed disease progression models should improve upon these statements.
3. Our simulations say that **complete removal of infections at child care, primary schools, workplaces and during leisure activities will *not* be enough to sufficiently slow down the infection dynamics**. It would have been better, but still not sufficient, if initiated earlier.
4. **Infections in public transport play an important role**. In the simulations shown later, removing infections in the public transport system reduces the infection speed and the height of the peak by approximately 20%. Evidently, this depends on the infection parameters, which are not well known. – This does *not* point to reducing public transport capacities as a reaction to the reduced demand, but rather use it for lower densities of passengers and thus reduced infection rates.
5. In our simulations, removal of infections at child care, primary schools, workplaces, leisure activities, and in public transport may barely have been sufficient to control the infection dynamics if implemented early on. Now according to our simulations it is too late for this, and (even) harsher measures will have to be initiated until possibly a return to such a restrictive, but still somewhat functional regime will again be possible.

Evidently, all of these results have to be taken with care. They are based on preliminary infection parameters taken from the literature, used inside a model that has more transport/movement details than all others that we are aware of but still not enough to describe all aspects of reality, and suffer from having to write computer code under time pressure. Optimally, they should be confirmed independently. Short of that, given current knowledge we believe that they provide justification for “complete lockdown” at the latest when about 10% of available hospital capacities for COVID-19 are in use (and possibly earlier; we are no experts of hospital capabilities).^1^

What was not investigated in detail in our simulations was contact tracing, i.e. tracking down the infection chains and moving all people along infection chains into quarantine. The case of Singapore has so far shown that this may be successful. Preliminary simulation of that tactic shows that it is difficult to implement for COVID-19, since the incubation time is rather long, people are contagious before they feel sick, or maybe never feel sufficiently sick at all. We will investigate in future work if and how contact tracing can be used together with a restrictive, but not totally locked down regime.

When opening up after lockdown, it would be important to know the true fraction of people who are already immune, since that would slow down the infection dynamics by itself. For Wuhan, the currently available numbers report that only about 0.1% of the population was infected, which would be very far away from “herd immunity”. However, there have been and still may be many unknown infections (Frankfurter Allgemeine Zeitung GmbH 2020).

## 2 Introduction

The SARS-CoV-2 virus is increasingly affecting the world’s population. Neither treatments nor vaccines are currently known. Although the infection does not lead to severe illness for many people, about 10-20% of infected persons need hospital care, and about 5% become critical (Robert Koch Institute 2020). Given, say, 22000 hospital beds (Berlin.de n.d.) for 3.5 million people in Berlin, it is clear that 100000 simultaneously infected persons, needing 10000 to 20000 hospital beds, would push the system beyond its limits. It is therefore plausible that the spread of the infection needs to be slowed down, to something like well below 10000 infections per week and ideally much less.

The general means for slowing down infections is well-known: distancing, and ultimately quarantine. If contagious persons do not meet susceptible persons anymore, then the virus cannot spread any further. Also, spreading does not necessarily have to be reduced to zero: Once every infected person infects less than one other person, the virus dies out. The general dynamics is captured in the so-called SIR model, with S = susceptible, I = infected, and R = recovered (Kermack, McKendrick, and Walker 1927; R. M. Anderson and May 1979). Every time a susceptible and an infected person meet, there is a probability that the susceptible person becomes infected. Some time after the infection, the person typically recovers.

The details of this process, however, are much more complicated. There is, for example, infection via the air (e.g. droplet infection), and infection by touch (smear infection). Smieszek (Smieszek 2009, 2010) has described in particular the first process in detail: Infected persons generate a “viral load” that they exhale or cough into the environment, and people close by have an exposure. Overall, the probability to become infected by this process in a time step *t* is described as

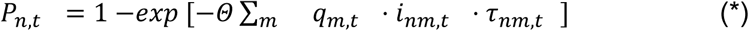

where *m* is a sum over all other persons, *q* is the shedding rate (∼microbial load), *i* contact intensity, and τ the duration of interaction between the two individuals. These microscopic parameters are difficult to obtain even for known virus species and can thus be used as calibration parameters. However, some mechanical aspects are clear:

- The contact intensity needs to go down with distance *d*. If one assumes that viral material from a point source is distributed homogeneously in all directions (similar to light), then *i* ∼ 1/*d*^*2*^. One would, however, rather expect a diffusive process, leading to *i* ∼ 1/*d*^3^. This implies that distance matters a lot; a distance of 1 meter instead of 1/2 meter would reduce the probability of contagion by a factor between 4 and 8. Even if this is only a rough approximation, this makes clear that one should account for typical distances between persons.
- The process is probabilistic. Even a close distance does not have to lead to an infection if it is short enough, and the shorter the time, the smaller the probability.

For SIR models to be interpretable, they need to be embedded into a contact graph (Bajardi et al. 2011; Skvortsov et al. 2007; Balcan et al. 2010). An infected person from out-of-country may return to its family, there it might infect one if her/his children, the child may take it to the school where it infects other children, they take it to their family, members of those other families may take it into public transport vehicles, to leisure activities, or to their workplace, etc. These social networks are typically not available for simulations, for example for privacy reasons.

It *is, however*, possible to generate synthetic approximations to these trajectories. This is routinely done in transport modelling. One approach is to use some information from mobile phone data (but not the full trajectories), and process them together with information about the transport system and with statistical information from other surveys (Senozon 2020b). That approach leads to synthetic movement trajectories for the complete population. From these trajectories, it is possible to extract how much time people spend with other people at activities or in (public transport) vehicles. It is also possible to estimate distances to other persons, e.g. by comparing the number of persons in a public transport vehicle with that vehicle’s size, or by making assumptions about such distances at typical activities (home vs work vs leisure vs shopping).

It turns out that such data sets are available for Germany, Switzerland, and Austria. This paper presents simulation results based on a cutout from that data set, for the case of Berlin. What is effectively done is to construct an SIR-like model on top of persons’ movement trajectories:

1. One or more infected persons are introduced into the population.
2. At some point, infected persons become contagious. From then on, every time they spend time together with some other person in a vehicle or at some activity, equation (*) is used to calculate the probability that the other person, if susceptible, can become infected. If infection happens, the other person will follow the same dynamics.
3. Contagious persons eventually become recovered.

The model runs many days, until no more infections occur. Note that compared with the SIR model we have differentiated the “infected” state into infected and contagious, in order to take account of the relatively long incubation times of COVID-19.

Now once such a model is available, containment strategies can be introduced into the model, and the results can be investigated. Optimally, the model should be calibrated before this is done. Smieszek et al (Smieszek 2009; Smieszek, Fiebig, and Scholz 2009; Smieszek et al. 2011) and Hackl/Dubernet (Hackl and Dubernet 2019, 2018) have done this for the influenza virus, and thus demonstrate that this is possible. Unfortunately, for the current situation calibration is not possible, since neither enough data about the microscopic behaviour of COVID-19 is available, nor enough epidemiologic data for model fitting. It is still plausible to assume that the structural responses of the model to distancing/quarantine measures remain valid, albeit quantitative results have to be taken with a grain of salt. We believe that it is better in the current situation to publish the results anyways.

The paper will consider the following containment strategies :

- No longer use public transport, i.e. people to go about their activities by individual means of transport (car, bicycle, walking).
- Shut down work activities, i.e. force people to either work from home or not work at all.
- Shut down leisure activities, i.e. force people to remain at home when they ordinarily would perform leisure activities.
- Combinations of the above measures.
- Complete lockdown, i.e. everybody stays at home.

The strategies will be introduced at different days during the infection dynamics.

The present paper goes beyond the above-mentioned studies (Smieszek 2010; Hackl and Dubernet 2019) in the following aspects:

- The present study includes the effect of public transport.
- The present study is based on existing real-world models, normally used for transport planning.
- The present study explicitly looks at distancing/quarantine measures.

A somewhat similar study to ours is by (Ferguson et al. 2020). They have considerably more experience in the area of epidemics modelling. However, we believe that our human movement and resulting contact model, based on real world data, goes beyond their work. We thus hope that our work will be a contribution, in particular for regional-scale modeling of epidemics and containment measures. The approaches and their results are compared in more detail in a discussion chapter at the end of the paper.

## 3 Details

### 3.1 Input data

As stated earlier, the input data stems from activity-based transport modelling (Hilgert et al. 2017; Axhausen 1989). Such models generate complete daily activity chains of persons, for example something like home–work–shop–home–leisure–home. Activities come with times and, importantly, locations. The activity chains are then used as input for (agent based) transport simulation models, which assign activity locations, modes and routes and thus generate emergent effects such as congestion and emissions (Horni, Nagel, and Axhausen 2016). The models are used, e.g., for forecasting effects of autonomous vehicles (Bischoff, Maciejewski, and Nagel 2017), investigating traffic signals (Thunig, Kühnel, and Nagel 2019), understanding and reducing emissions (Kickhöfer and Nagel 2016) or noise (Kaddoura, Kröger, and Nagel 2017) effects of transport, or evaluating strategies for carbon-reduced or carbon-free transport (Nagel et al. in preparation).

The input data for the present project is generated by Senozon (Senozon 2020b, [a] 2020), and has been used by us also in other projects (VSP 2020). For the present paper, the activity chains are just used “as is”. From their information, it is possible to reconstruct the time-dependent trajectories of each person with respect to their “containers” (facilities or vehicles), and it is possible to reconstruct who shares, at what time, which container (cf. Fig. 1).

**Fig. 1.**
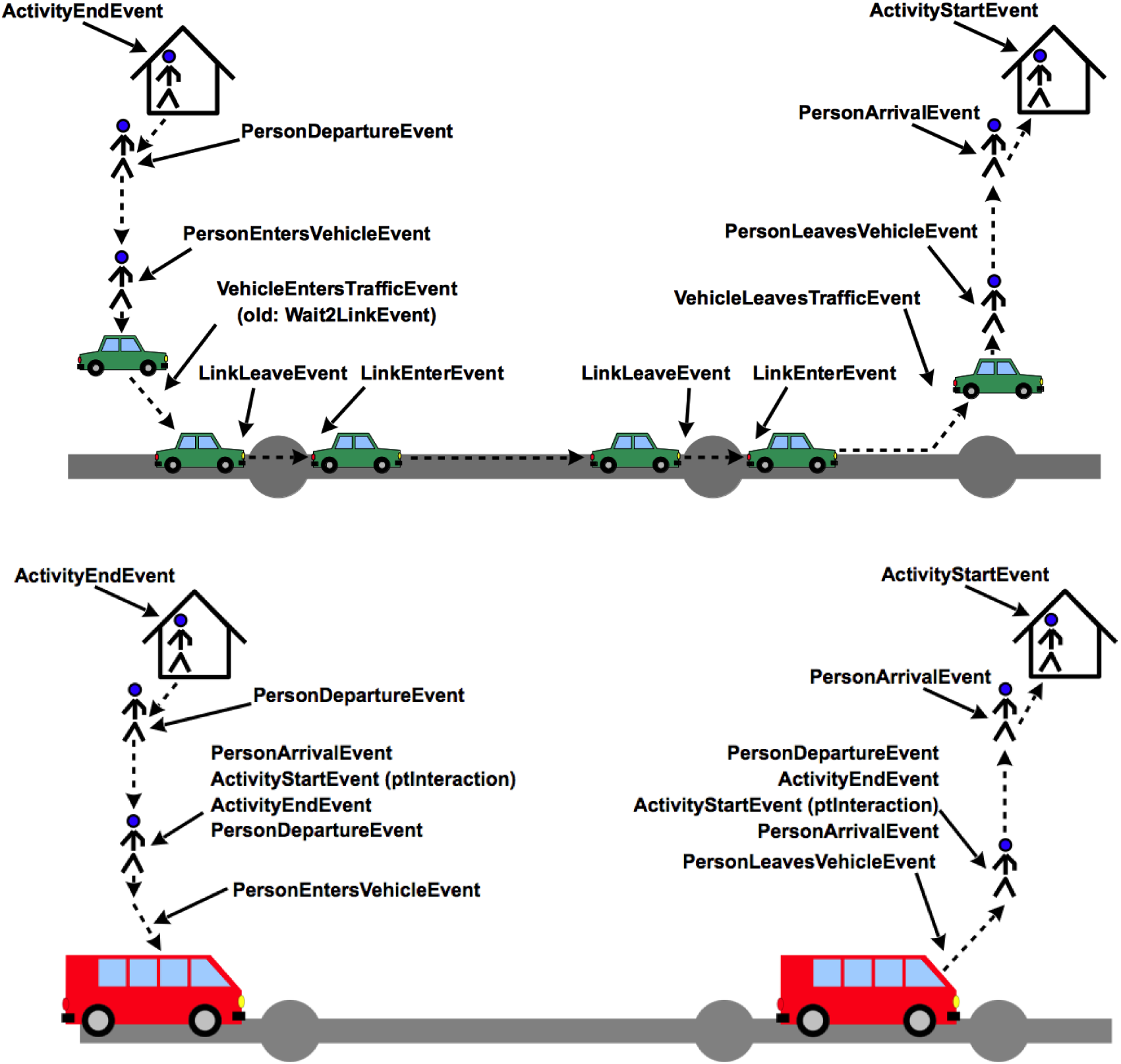
Top: Events for travel by individual vehicle. Bottom: Events for travel by public transport.

### 3.2 State transitions

The model for the infection itself is described in the next section; this section will first explain the possible agent states. Once a synthetic person becomes infected on day number *d*, it will undergo the following state transitions (Roy M. Anderson et al. 2020):

- Between days d+3 and d+4, it will transition to *contagious*. Only then will it start infecting other synthetic persons.
- Between days d+5 and d+6, it will self-*quarantine* with a probability of 20%. This is modelling that only 20% of people show strong enough symptoms to recognize that they are sick, and at which point they withdraw from their activities (Frankfurter Allgemeine Zeitung GmbH 2020). The current model assumes that they go into full quarantine, i.e. they also do not infect people at home.
- Between days d+15 and d+16, it will transition to *recovered*. At this point, it is assumed that they resume their activities, but that they are immune (Bao et al. 2020).

As one sees, this is a somewhat more elaborate version of the standard SIR (susceptible – infected – recovered) model, to take into account the peculiarities of Covid-19 as far as they are currently understood (Roy M. Anderson et al. 2020). State transitions always happen in between two runs of the daily dynamics (see below) – that is the reason why we say “between days d+x and d+x+1”.

### 3.3 Infections at facilities and in vehicles

The algorithm looks at agents when they *leave* a facility. At this point, it

- randomly selects 3 other agents which are at the facility at the same time, and
- with each agent computes a possible infection if either the leaving agent is susceptible and the other agent is contagious, or the other way round. The infection model is equation (*), the time τ is the time (duration) that both agents were simultaneously at the same facility.

We bound the infection dynamics at 3 other agents, since we assume that persons do not interact with everybody in the facility. If there are fewer than 3 other agents at the facility, then interaction simply happens with everybody. Since we are using a 25% scenario, the number “3” stands for 12 other persons with which interactions happen; this seems to be a (somewhat) plausible number for interactions at workplaces. Clearly, we do not want the model to interact with everybody at the facility.

The same algorithm is used for interaction in vehicles. In terms of implementation, it uses a generalized dynamics for *containers*, and treats both facilities and vehicles as such containers.

Note that the number “3” (or implicitly “12”) is used when the agent leaves. There is, however, also interaction when other agents leave. Thus, this rather models the interaction with “available spaces” than with persons. E.g. assume the following time line (from left to right) and consider in particular the agent X:

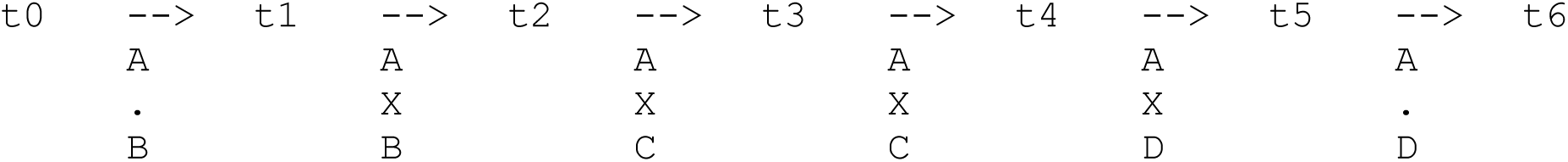

This can, for the ease of interpretation, be interpreted as three seats in a row in a public transport vehicle. We have the following events with respect to person X:

- At time t1, X enters the vehicle, and takes a seat between A and B.
- At time t2, B leaves the vehicle. In consequence, a possible infection of X from B is computed with equation (*), with t2 - t1 as the duration of the interaction. Also, C enters the vehicle, and takes the seat of B.
- At time t4, C leaves the vehicle. A possible infection of X from C is computed, with t4 - t2 as the interaction time. At the same time, D enters the vehicle, and takes the seat of C.
- At time t5, X leaves the vehicle. A possible infection of X from A is computed, with t5 - t1 as interaction time, and from D, with t5 - t4 as interaction time.

In the algorithm, however, we do not explicitly model seating positions, but just assume that every synthetic person that leaves interacts with up to 3 randomly selected other agents.

The variation over time of the density of persons inside the container is currently not taken into account. This could, however, be done in a further modelling step, given data of facility and vehicle sizes. What *is*, however, taken into account is the thinning out of persons in the container when they are in quarantine: The algorithm still computes interaction with up to 3 randomly selected persons, but if these persons are in quarantine, then there is no infection dynamics in either direction.

### 3.4 Multi-day modelling

Optimally, one would have multi-day trajectories. In our case, the data that we have ends at the end of the day. Our simulations thus run the same person trajectories again and again. This presumably *underestimates* mixing (see next section). However, there is still strong mixing because the synthetic persons interact with other persons at the same facility every day. For example, a public transport train may have 1000 passengers. In our 25% scenario, they would be represented by 250 passenger agents. Out of these, any of our agents would only see 3 other agents representing 12 persons. This leads to different encounters in every synthetic day, even when repeating the same trajectory over and over.

The same holds for facilities, where the data is actually constructed such that there are typically around 400 persons per facility. That is, any work, shopping, or leisure facility can have up to 400 visitors at the same time, and every person that has that facility in its trajectory can interact with all of these persons. This number of interacting agents is, however, limited (see infections at facilities and in vehicles). This results in an issue for home locations: Our data model does not differentiate between “persons living in the same block” and “persons living in the same household”. With the current state of our modelling, the infection probability at home locations thus will have to average over these situations.

During the first simulated day 10 random agents are infected. The first iteration is then used to construct certain data structures meaning that every agent memorizes its trajectory. Agents are put in and removed from the respective containers when undergoing activity start and end events and vehicle enter and leave events. These facility containers are not cleared at the beginning or end of the day meaning that agents can remain in a container from one day to the next. This is important to calculate the duration times accurately for the infection dynamics - especially at home facilities. Some agents, however, do not have closed circles, meaning that their last activity is not the same one as their first one. In these cases it is assumed that agents start their first activity at 00:00.

As stated before, activities are of a certain type. These types are used in this research to determine the effects certain strategies have on the epidemic spreading. If an activity type is closed, agents will still be put in the respective containers without taking part in the infections dynamics. This is important especially for the dynamics in transit vehicles, because agents travelling to closed activities essentially represent “holes” as they would not travel at all in reality.

### 3.5 Graph interpretation

The following borrows from social network and containment studies done by many others (Brockmann, David, and Gallardo 2009; Makinde 2007; Bajardi et al. 2011; Colizza and Vespignani 2008; Colizza et al. 2007). It is re-interpreted for our specific situation.

The travel model can be seen as generating an interaction graph. In a first step, one could connect persons to their activity facilities, e.g. all persons working at a certain facility would be connected to it. In a second step, one could remove the intermediate facility from the graph, and draw direct connections between the persons. These connections include a time period, e.g. person A and person B were simultaneously at facility W between time1 and time2. The same applies to public transport, i.e. persons C and D may have been simultaneously in public transport vehicle V between time3 and time4.

As stated above, being at the same facility does not necessarily imply immediate interaction. According to the models described earlier, there is a certain probability to interact, and there is mixing from one day to the next. In terms of the graph interpretation, the model thus implies weak links between persons who are in the same vehicle or at the same facility if these containers are populated with many persons at that time.

It is now easy to see that virus progression would quickly die out if everybody remained at home: Then each home facility would generate a subgraph (= a clique, since everybody in the subgraph would be directly connected to everybody else); the infection would continue to spread in those cliques that have at least one infected person when this type of quarantine is started; and that would be it.

If we now assume that persons do *only one* other activity, such as work, then the virus can jump from one such clique to another via the workplace. It is not clear if this would infect the whole population or not, since there might be subgroups of cliques that are strongly connected, but the subgroups may be sufficiently disconnected that the virus cannot jump. Evidently, all other activities generate further interactions with yet other cliques, and public transport vehicles will do the same.

For an example, look at Fig. 2. There are four families, with 4, 4, 3, 1 member(s). Family 1 has one member going to “work1”, and one other member going to “school”. Family 2 has one member going to the same “school”, and one other member going to another workplace “work2”. Family 3 has one member going to “work2”, and using a certain “pt vehicle” (presumably on the way to and from work). Family 4 consists of only one member, which uses the same “pt vehicle”, but goes to yet another workplace “work3”.

**Fig. 2.**
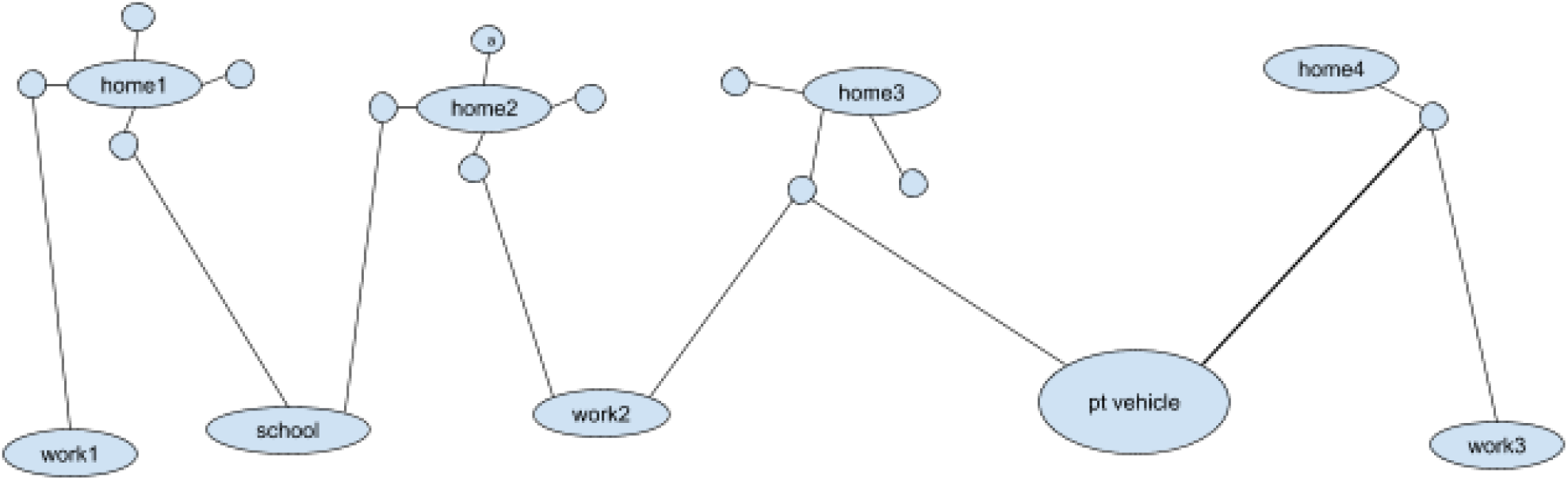
Example of graph connections between persons and containers (facilities and vehicles).

Now assume that initially only person “a”, in “home2”, is infected. One has the following cases:

- If everybody stays at home, then all other members of “home2” will become infected, and then the virus will travel no further.
- If home and work are open but school and public transport are closed, then in the example the virus will also travel to “home3” and all its members, but then stop. This, however, depends on families 2 and 3 visiting no second work location, either by the same person or by another member of the families.
- If home and work are open and public transport can be used, then in the example the virus will also travel to “home4”.

That is, the connectedness of the graph decides about how far the virus will travel.

A second element is the infection probability. As indicated by equation (*), it is a result of microbial load, contact intensity, and contact time. In consequence, an interaction at work, with physical distance to co-workers but much time spent together, can have the same infection probability as a short time in a public transit vehicle while squeezed together.

A third element comes from the day-to-day repetition, as explained earlier. At home, one interacts with the same persons over and over again, effectively increasing the infection probability ever more, implying that either all members of the household are infected, or none. This results in thick lines connecting household members. *Households do not provide mixing beyond themselves*.

Public transport, however, is much different: One sits or stands next to other persons every day. In consequence, possible infection paths are between different persons every day. Since they run back into their respective homes, *public transport results in strong mixing*. Other activities are probably in between: children at day car or primary school are probably strongly connected within relatively small groups, i.e. similar to the infection dynamics at homes, while young adults at secondary or tertiary education are probably much less strongly connected, but have many possible interactions with other people, varying from one day to the next. For work it presumably depends on the work environment: workers in a small company meet the same people every day while workers in a large company may meet different people every day. People with customer-facing jobs, such as sales people, are again different.

This graph interpretation will be used in the following to provide possible intuition for the simulation results.

### 3.6 Calibration

Most parameters of the model are taken from the literature, as explained so far. The remaining free parameters are, from equation (*), Θ *q i*, where *q* was the shedding rate, *i* the contact intensity, and Θ is a calibration parameter. Since none of these numbers seem to be known for COVID-19, we have proceeded as follows:

- We have set the base value of *q i* to 1.
- For public transport vehicles, we have set *q i* to 10.

As stated earlier, the increased contact intensity value for public transport models includes that distances between persons in public transit vehicles are often much smaller than in most other situations. Clearly, this is debatable, and simulations with other parameters could be run. Our intuition, however, is that a ratio of 10 in contact intensities between “transit vehicle” on the one hand, and “shopping”/”work”/”errands”/”(higher) education” on the other hand, is plausible. In contrast, for home and leisure we would expect higher values than those that we are currently using.

The remaining Θ parameter was then calibrated so that in the base case, the growth rate is a tenfold increase of infected persons per 7 days. This is larger than what we see now in Germany (where we have a tenfold increase per 9 days), but is a plausible fit to the initial days which were totally without distancing measures, which is what our base case represents.

## 4 Runs and results

### 4.1 Base case

Fig. 4 shows the base case simulation. The base case assumes no reaction at all meaning that people behave completely normal and no containment strategy is in place. This behaviour is of course improbable when faced with an epidemic with such severe proportions. The base case simulation does, however, include that 20% of the infected persons self-quarantaine, similar to staying at home when sick.

**Fig. 3.**
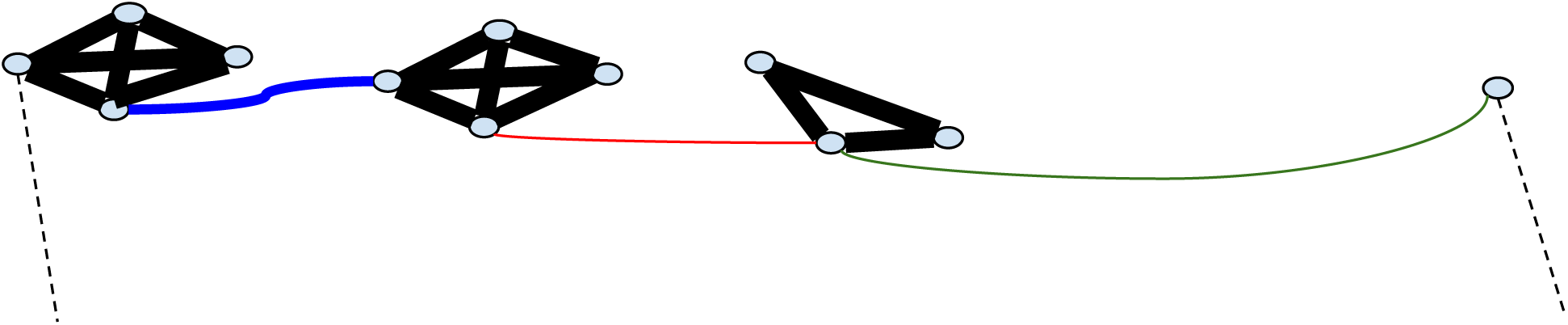
The interaction network from figure 2 when the intermediate containers are removed.

**Fig. 4.**
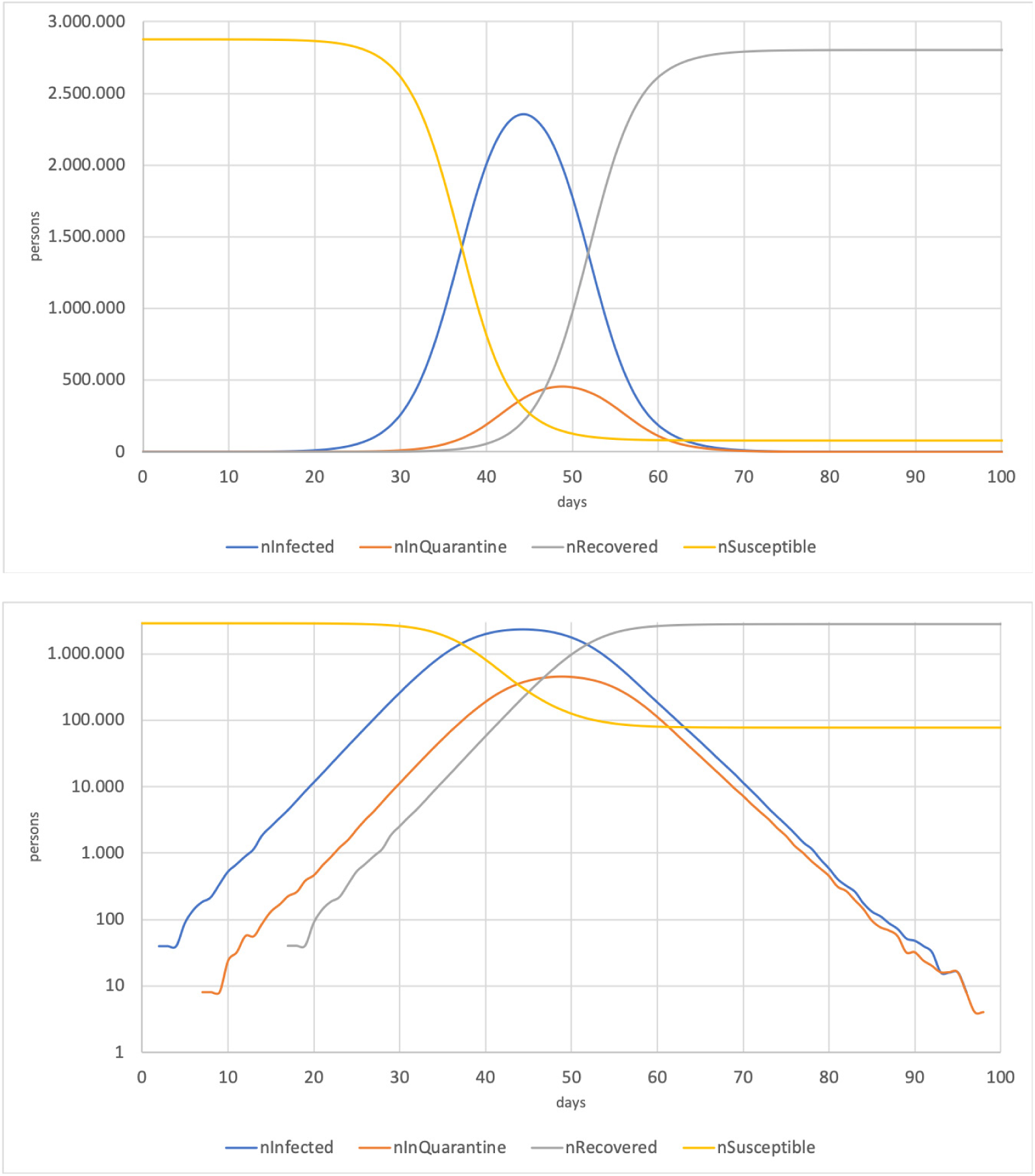
Top: Base case (linear). Bottom: Base case (log).

One notices the following aspects:

- There is exponential growth of the number of infected persons initially. This also means that in the first approximately 10 days the number of infected persons is small, and the situation does not seem dramatic. This then rapidly changes, eventually leading to about 2.3 million simultaneously infected persons around day 44. Evidently, this is what we should not let happen; according to the above numbers it would imply 230,000 persons needing hospital care, many times more than what the system can absorb.^2^
- The number of recovered persons goes up to about 97% of the population.^3^ Once that many persons are immune, the reinfection rate drops below 1, and the virus dies out.
- The model does not explicitly contain death cases. For the dynamics of the model, it is irrelevant if infected persons eventually recover or eventually die.

Fig. 5 shows the share of infections occurring in different containers. The outcome shows that especially home, leisure, work and public transit activities have a high influence on the epidemic spreading. As the home activity cannot be reduced, the next sections will outline the effects of reducing leisure, work and public transport. One notices that about a fourth of infections occur in public transit vehicles. Our current assumption is that the contact intensity *ii*, as described earlier, is 10 times higher in public transport vehicles than in activities.

**Fig. 5:**
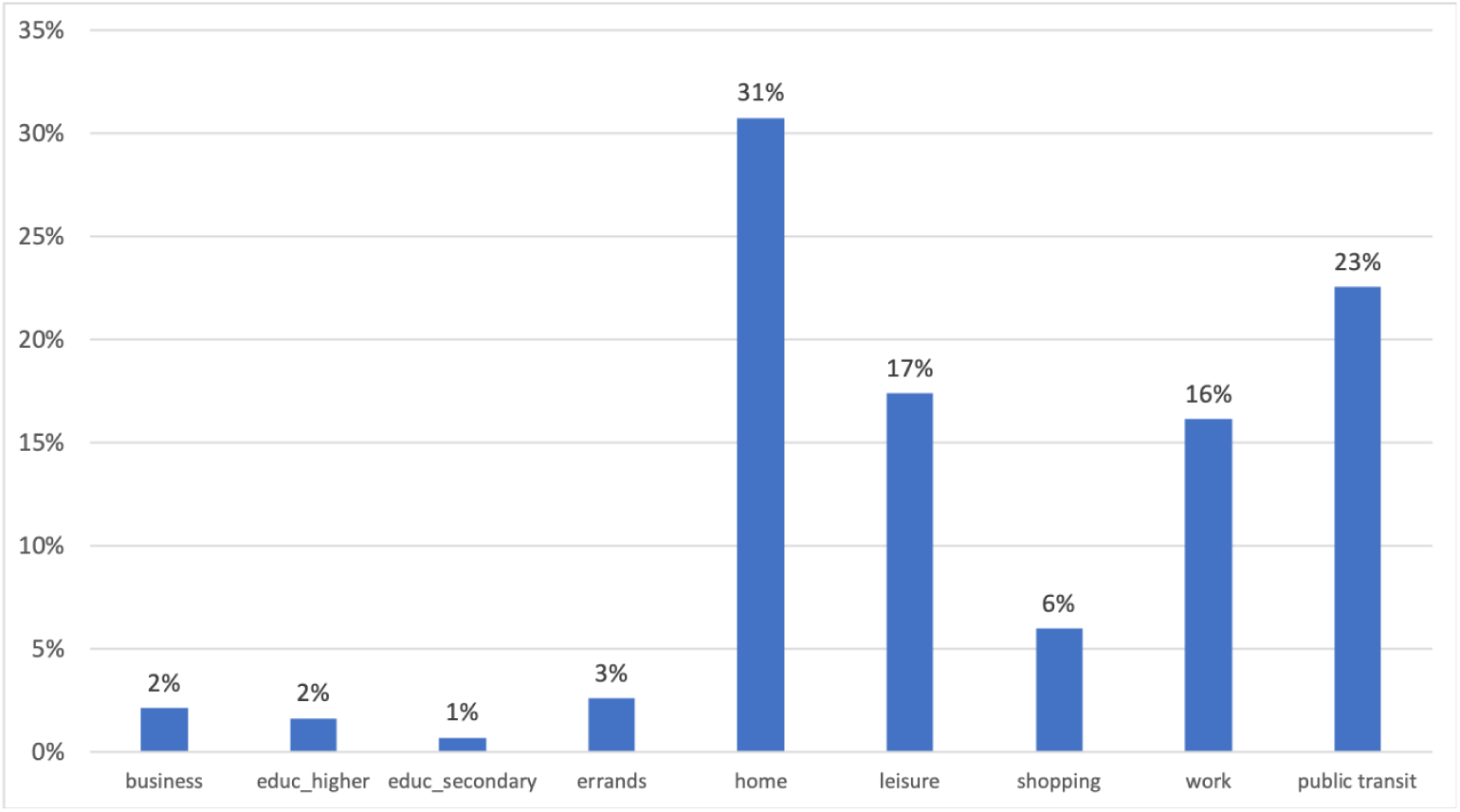
Distribution of infection types in base case.

### 4.2 Assuming no infections in public transport

Fig. 6 shows the effect a complete shutdown of public transport would have. The cases differ from each other with regard to the time point the measure is introduced. The maximum number of simultaneously infected persons goes down in every case, and the maximum is reached later compared to the base case. Also, the overall number of infected persons becomes smaller, implying a lower threshold of herd immunity. It becomes clear that shutting down public transport has an impact even when enforcing it as late as day 30.

**Fig. 6.**
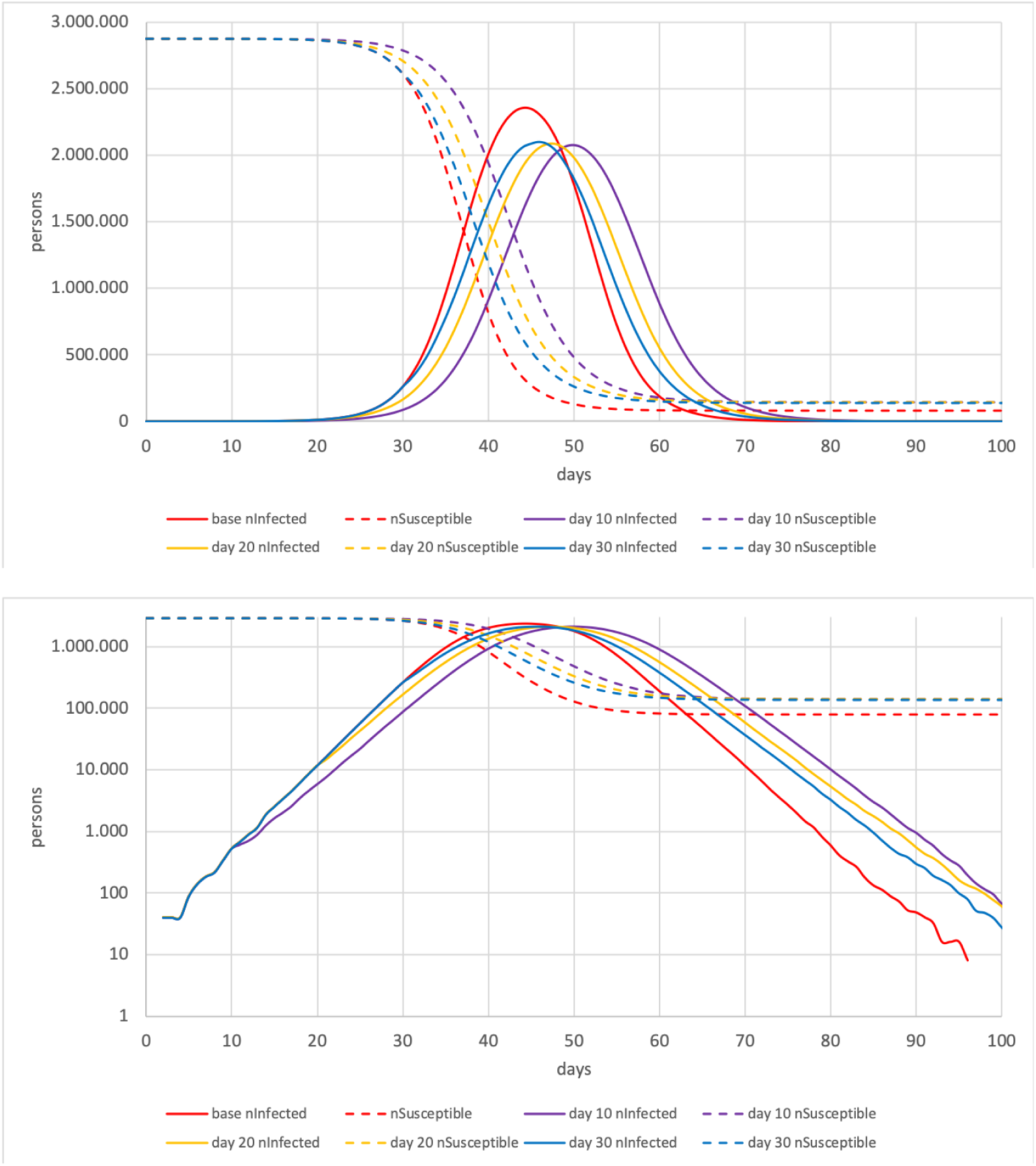
Top: No public transport cases (linear). Bottom: No public transport cases (log).

Note that we are not recommending to shut down public transport. Still, the contagion effect of public transport clearly plays a role. Resulting recommendations include:

- Public transport should run at the highest frequency, with the largest buses and the longest trains possible, so that people can spread out, and in consequence the contact intensity becomes smaller.
- Public transport vehicle drivers should be protected, since otherwise running at high frequencies will eventually no longer be possible because of lack of drivers.
- The authorities need to consider reimbursing the operator for running large capacities despite reduced demand and thus reduced fare income.

### 4.3 Assuming no infections at the workplace

Fig. 7 shows the effect of no infections at the workplace. Again, the influence of the enforcement time point is shown. Shutting down the workplace has a severe effect as the maximum number of simultaneously and overall infected persons is significantly reduced in all cases compared to the base. If introduced no later than day 20 the peak also occurs later. This containment strategy also has a significant effect because infections in public transport to and from work activities no longer occur. Even though the number of simultaneously infected persons is reduced the numbers are still too high for our health system to cope with. Resulting recommendations include:

**Fig. 7.**
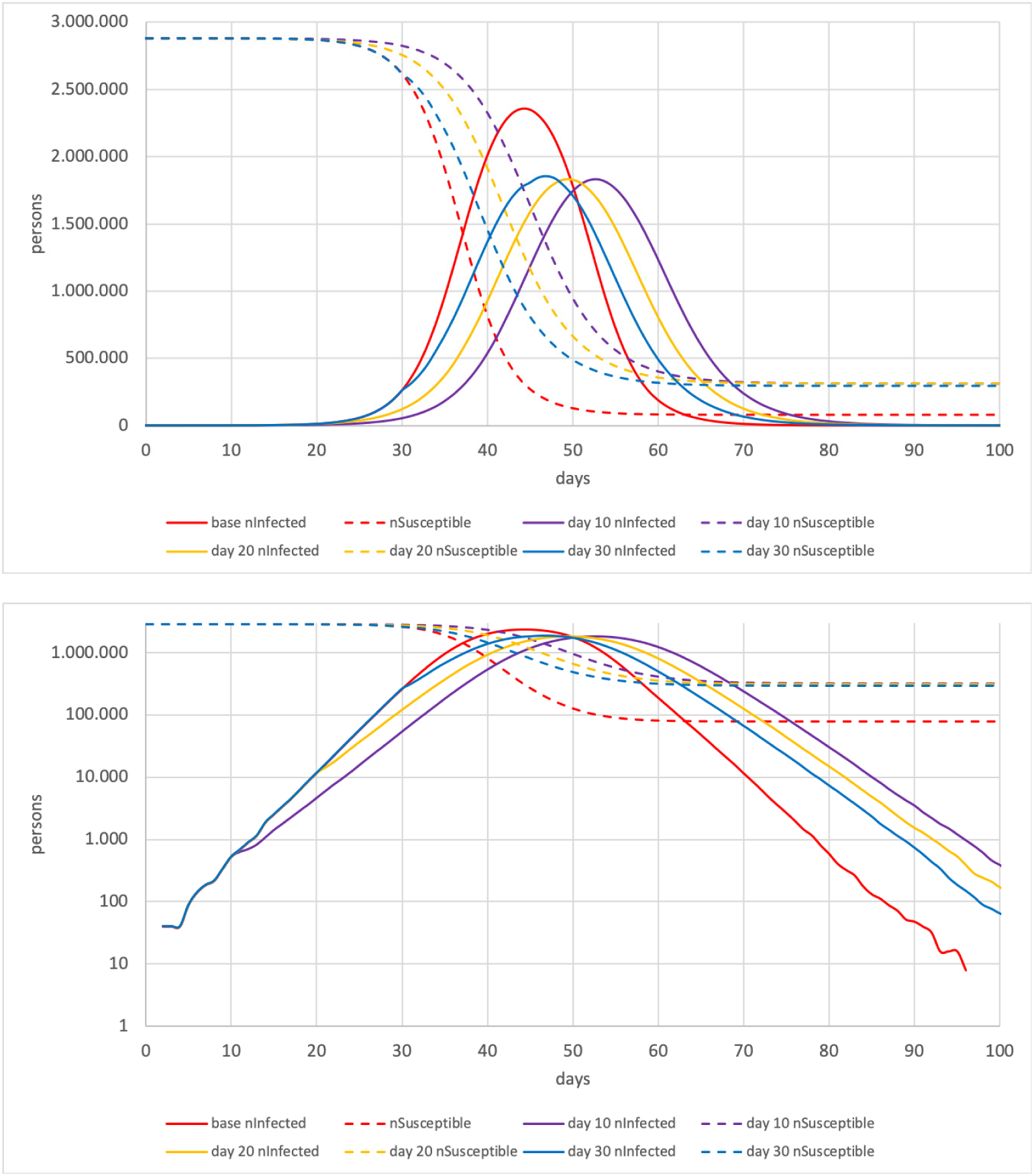
Top: No work cases (linear). Bottom: No work cases (log).

- It is of course not realistic to shut down all work activities. However, work activities should be reduced to a minimum to reduce the number interpersonal contacts meaning that people should work from home when possible.
- This strategy should commence early enough to shift the peak to a later time.

### 4.4 Assuming no infections at leisure activities

Fig. 8 shows the effect of no infections at leisure activities. The influence the enforcement time point has is shown. These measures noticeably reduce the peak and overall number of infected persons compared to the base. They shift the maximum to a later time when enforced before day 20. The following is recommended:

**Fig. 8.**
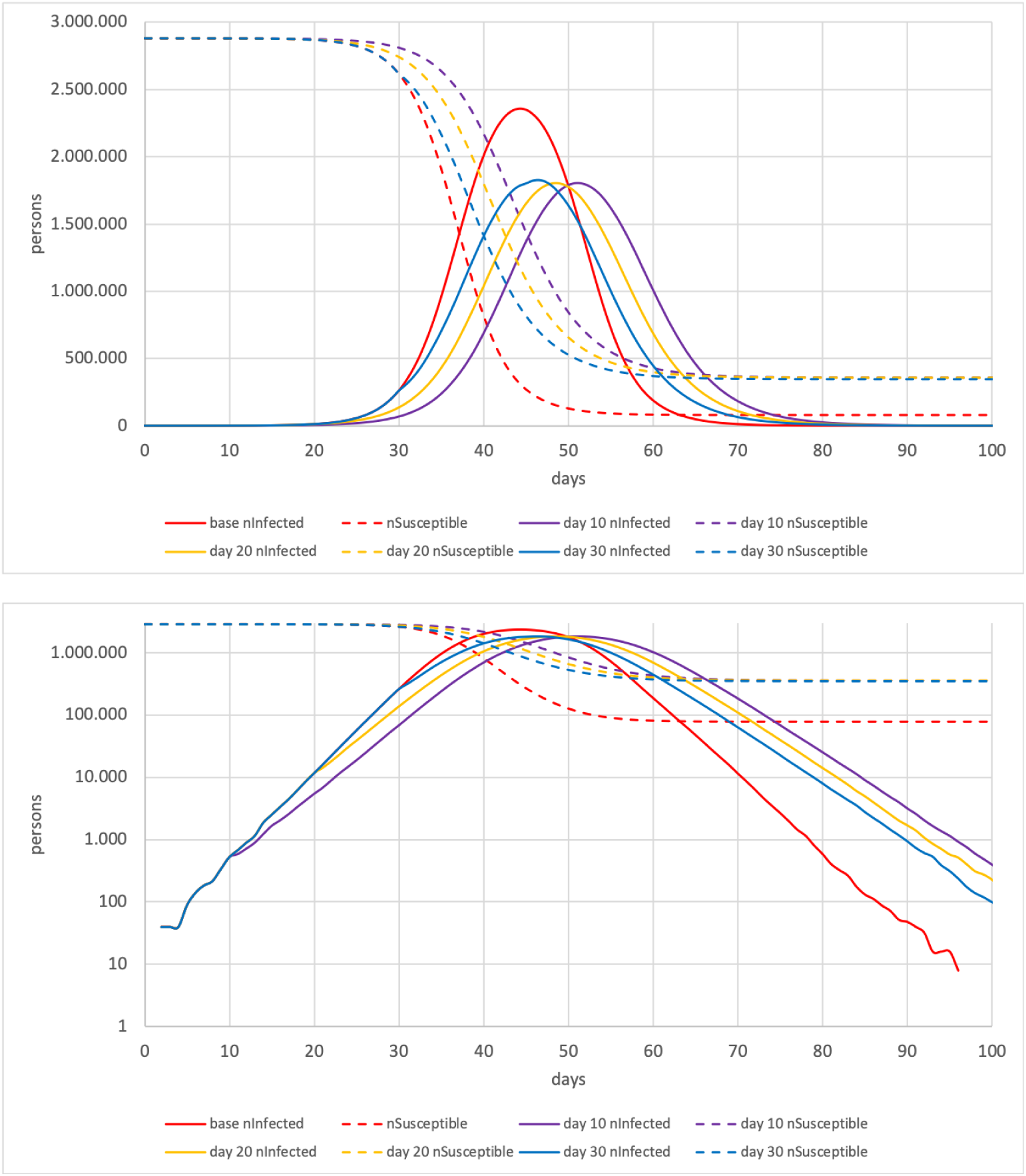
Top: No leisure cases (linear). Bottom: No leisure cases (log).

- Leisure activities should no longer take place as they increase both the speed and the impact of the epidemic.
- This restriction should be enforced as early as possible to shift the peak to a late time point.

### 4.5 Assuming no infections at work and leisure activities

Fig. 9 shows the impact of combining the containment strategies of preventing infections both at leisure and workplace activities. Compared to the single strategies it can clearly be seen that a combination of the two has a much stronger effect. The peak number of simultaneously infected persons can be reduced to about 970.000 cases when enforcing this strategy no later than day 20. This underlines the recommendations stated before that work and leisure activities should be reduced early enough. However, 970.000 simultaneous cases are still too many for our current hospital system to process, even though it is a very drastic measure.

**Fig. 9.**
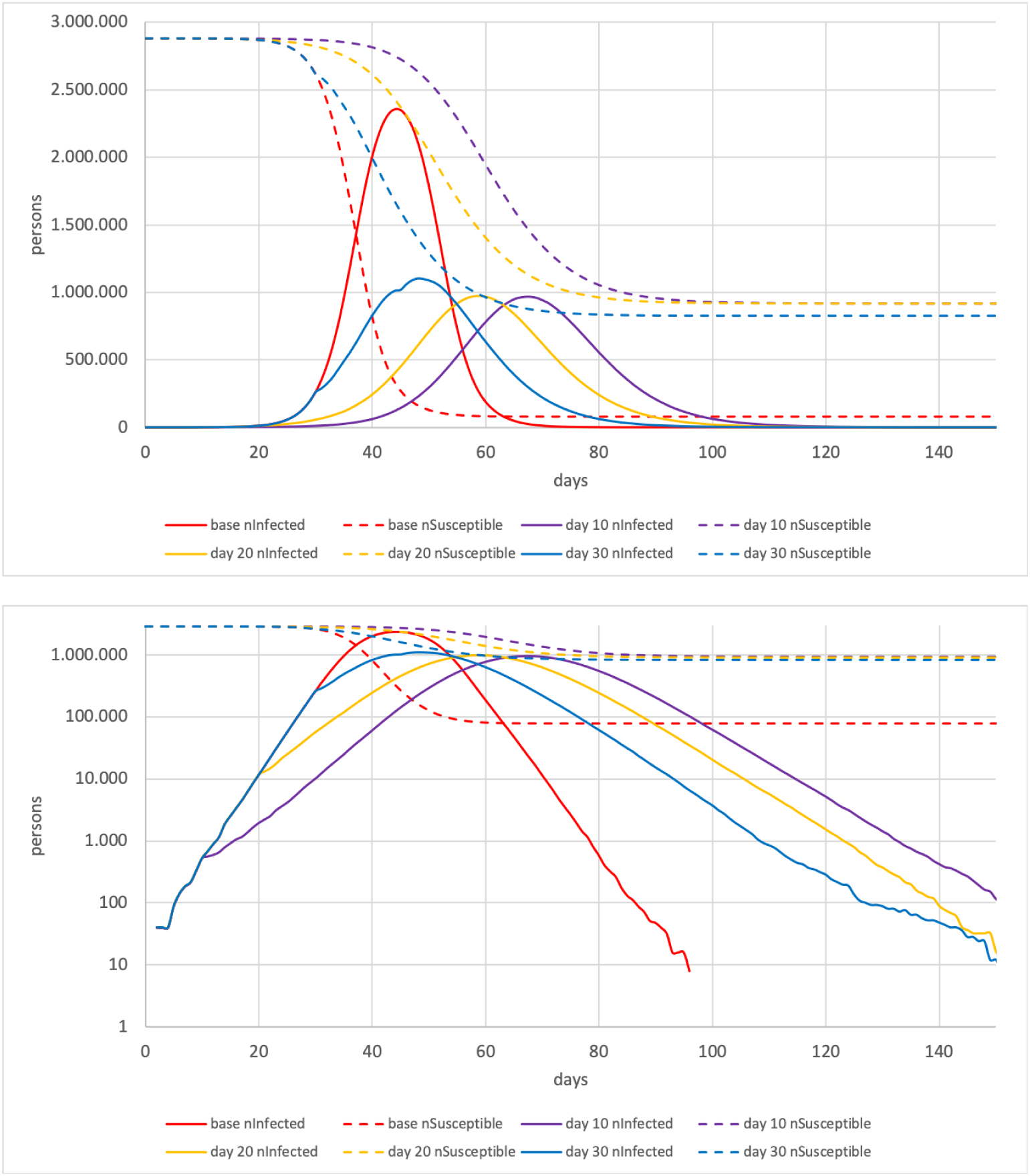
Top: No work and leisure cases (linear). Bottom: No work and leisure cases (log).

### 4.6 Assuming no infections at work, at leisure activities, and in public transport

Fig. 10 shows the effect of closing both work and leisure activities and also shutting down public transport. Compared to the section before, the maximum number of simultaneously infected persons can be reduced even more to approximately 880.000 persons when enforcing this strategy no later than day 20. As stated before, we do not recommend closing down public transport. However, this analysis shows that the population should reduce unnecessary trips where possible.

**Fig. 10.**
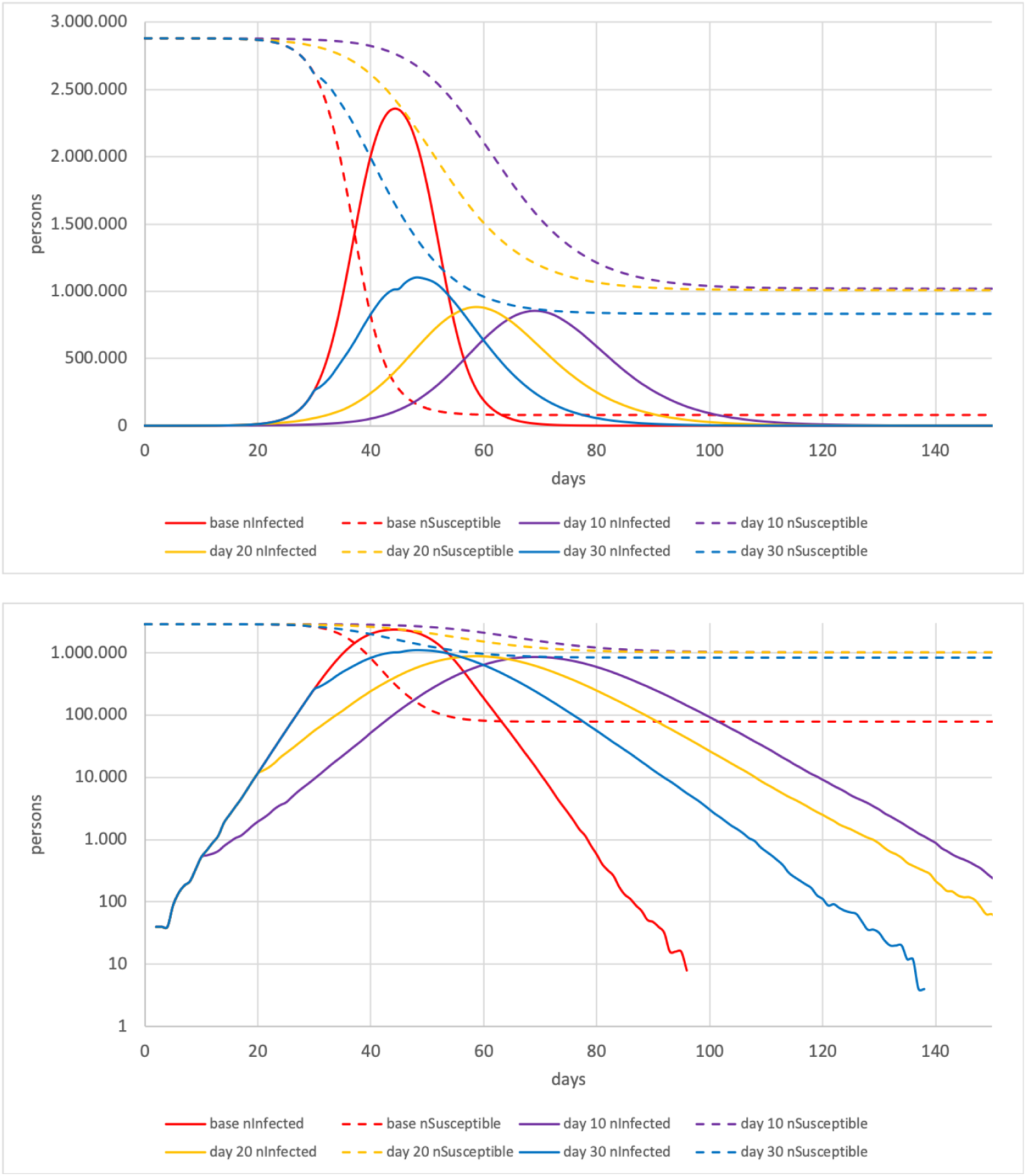
Top: No work, leisure and public transport cases (linear). Bottom: No work, leisure and public transport cases (log).

### 4.7 Complete lockdown

Fig. 11 shows the influence of complete lockdown. We define a complete lockdown as everyone stays at home. There are no other activity types any more. It can clearly be seen that a complete lockdown is effective shortly after enforcement. Even when done after 40 days it still helps to quickly reduce the number of infected persons. The log plot states clearly that complete lockdown stops the exponential growth of infections. This is of course a very drastic method which could be enforced when it becomes apparent that hospitals will become overloaded.

**Fig. 11.**
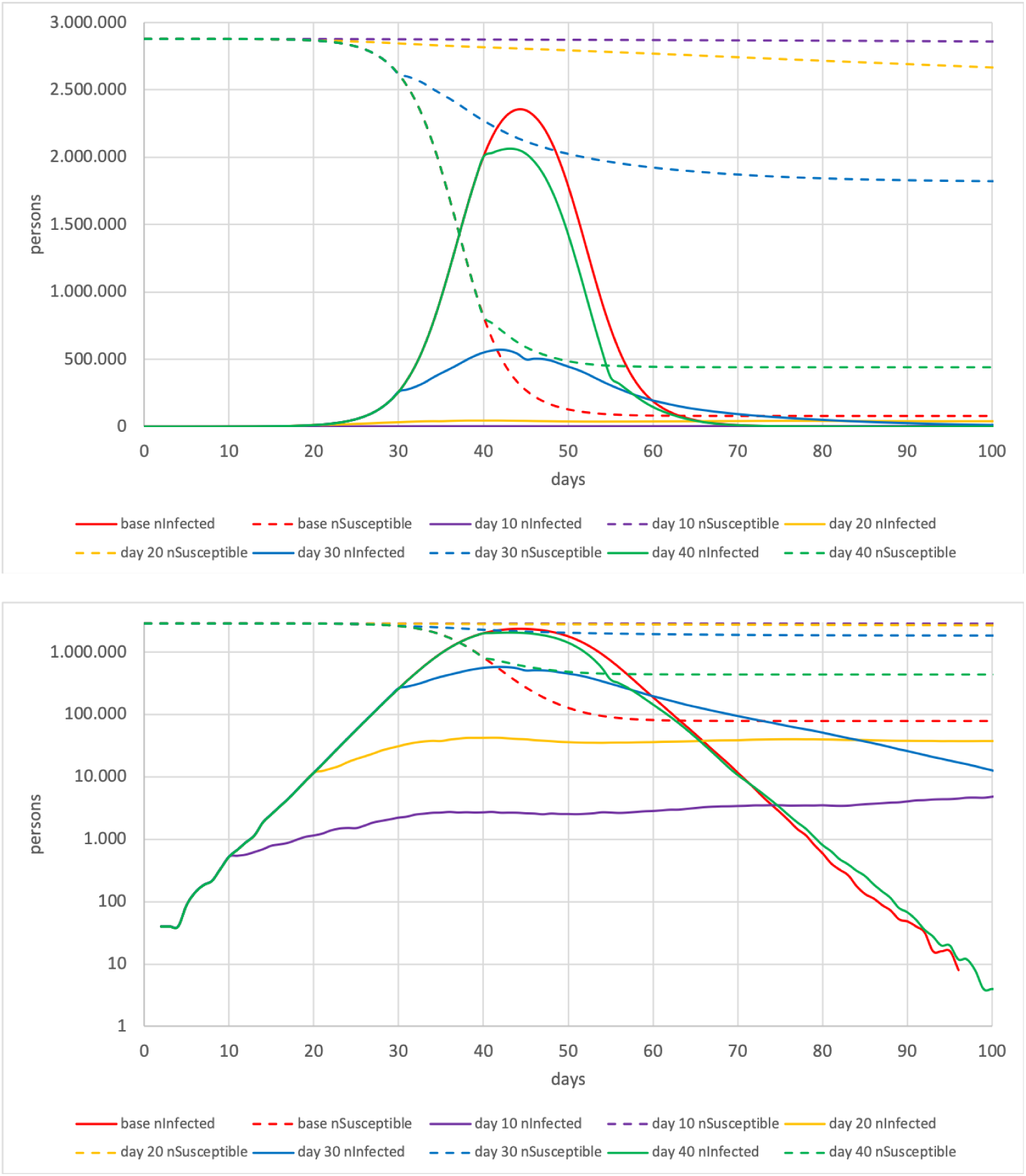
Top: Shutdown cases (linear). Bottom: Shutdown cases (log).

For many reasons, lockdown in reality will never be as complete as lockdown in our simulation model. Still, the experience from Wuhan seems to imply that it will bring the desired results for COVID-19. In Italy, it seems that people can still go to work, which may explain why, with respect to infection containment, the measures do not work as well as one might hope.

### 4.8 Comparison of containment strategies

All cases show that after enforcing a containment strategy one notices that, in the log plots, the slope of the infections immediately switches to a slower slope. This is especially the case when shutting down work and leisure activities, and with a complete lockdown.

We think that this implies that the statement “measures need to be implemented early” has to be differentiated:

- The rate (= slope in log plot) of the *infections* can be influenced at arbitrary points in time, and it follows the new policy measures immediately.
- Evidently, all infections that have been started before will still progress, and thus the slowdown of the number of critical cases will need the corresponding number of days to show in the data. For COVID-19, this time delay may be a week or more.
- The maximum number of simultaneously infected persons *is* influenced by how early or late the measure is implemented. A very plausible explanation is that, if strong separation measures are implemented early, this leaves many subgroups without a single infected person in that subgroup. If implemented later, the infection will already have spread to many of these subgroups.

Fig. 12 compares the maximum number of simultaneously infected persons in the different containment strategies. It can clearly be seen that solely shutting down public transport, work or leisure activities is not sufficient to reduce the number of patients needing hospital care simultaneously. The statement that single measure strategies may not be sufficient is consistent with results from other studies (Ferguson et al. 2020). The figure shows that rather drastic measures seem to be necessary:

**Fig. 12.**
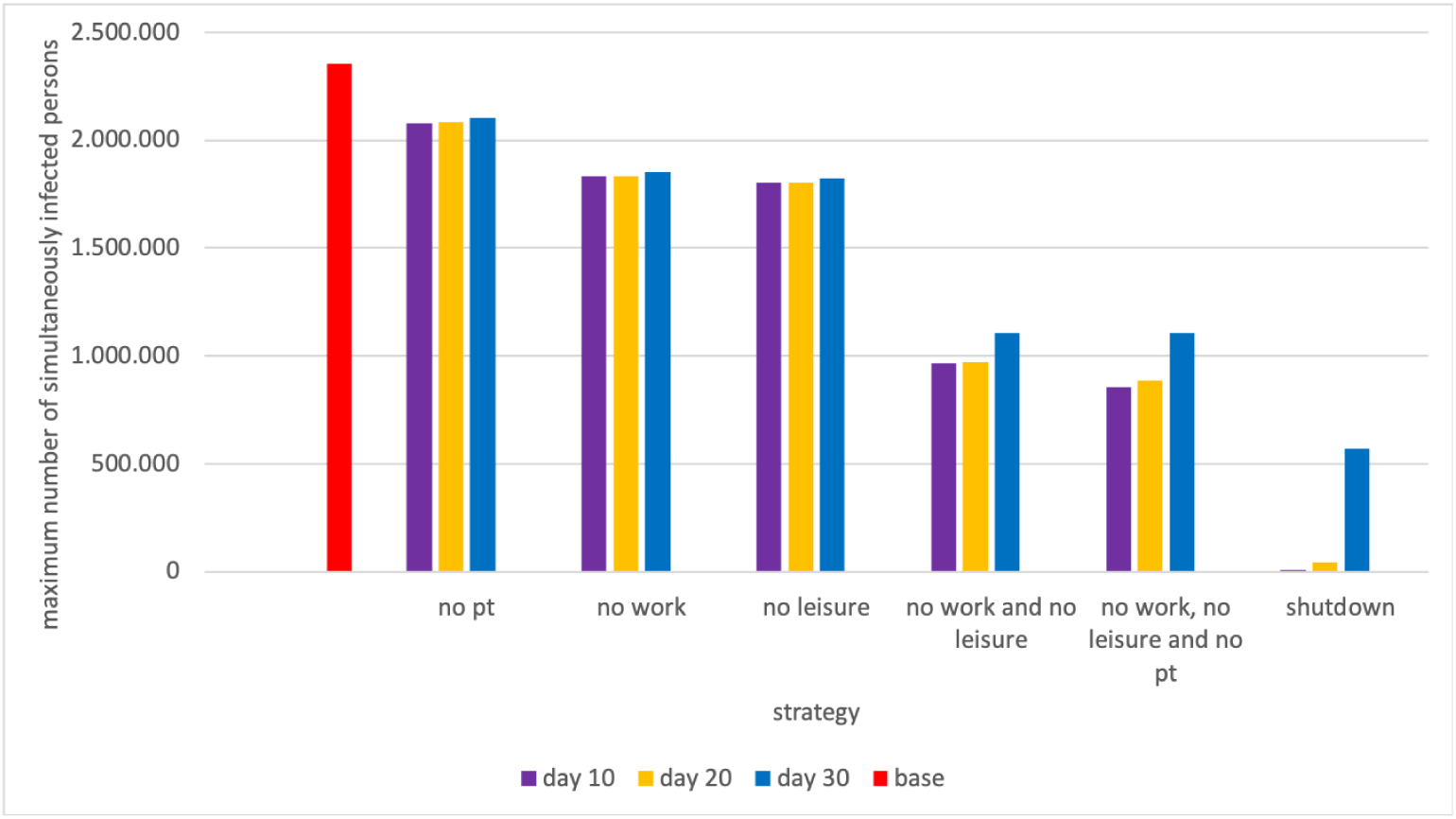
Comparison of maximum simultaneously infected persons.

- Closing leisure and work activities leads to the number of simultaneously infected persons being reduced. However, the reduction is, presumably, not sufficient.
- A complete lockdown would lead to a sufficiently low number of simultaneously infected patients if done in time.
- If such drastic measures are not enforced, the number of simultaneously infected persons would probably lead to high numbers of hospitalized persons. This would make a high hospital capacity necessary.

## 5 Discussion

None of the measures simulated in this paper, short of complete lockdown, is sufficient to reduce the height of the peak to levels to where we assume that they can be handled by the health system. Presumably, more drastic measures, as they were undertaken in China and are currently undertaken in Italy, will also have to be implemented in Berlin (and Italy’s strategy of still allowing persons to go to work may prove insufficient).

Assuming that these more drastic measures work, one also has to assume that they will have to remain in place for a rather long time, and can only very slowly be relaxed, together with the increasing share of immune persons in the population.

Our results are roughly in line with the work of (Ferguson et al. 2020), which appeared while we were working on our studies. Similarities include:

- If left to itself, the peak of the infection wave will generate numbers of critically ill patients that will far exceed existing hospital capacities.
- No single “simple” restriction will by itself be sufficient to bring infection numbers down to what hospitals can handle. A combination of restrictions will be necessary.
- Any combination of strategies that avoids over-utilization of the hospital systems will have to be in place for many months.

Ferguson et al. recommend that restrictive measures should be triggered by certain COVID19 loads of the hospital system, taking the time-delays of the infection dynamics into account. We agree with this assessment.

There are differences in the details. Ferguson et al. rely on an epidemics model that has been developed over many years and tested against other viral outbreaks, in particular influenza. Also, they include a relatively detailed progression model from infections to critical cases, which we omit. They also include spatially resolved demographics (which allows to identify shares of the particularly vulnerable elderly), which we could include in principle, but have so far not used.

On the other hand, we believe that our human movement model is better: Rather than randomly assigning workplaces, we use a data-driven process (which, however, is still synthetic because of privacy constraints). Furthermore, we include *all* activities of persons, again, because our model is data driven. Even if our persons are synthetic, their activities come from upstream data. Also, we include the effects of public transport. Finally, we include a mechanistic infection model, taken from Smieszek (2010). Ferguson et al. (2020) do not describe their microscopic infection model in that paper; we cannot tell if the reported so-called “R” values (= reinfection values) are model input or model output.

Ferguson et al report a later peak than we do. The causes of this should be clarified. They presumably include:

- Ferguson et al report critical hospital cases, while we show infected cases. Clearly, hospital cases show up later than infected cases.
- The model of Ferguson et al runs for the whole country (of Great Britain), while we run the simulation for Berlin. It is plausible that the infection progresses more rapidly in a relatively homogeneous and well-connected city like Berlin than across a whole country which includes many more remote regions.
- We may have calibrated against different growth rates. The paper by Ferguson does not contain a logarithmic plot. Taking it from a linear plot, they seem to have a doubling of cases every 10 days. This is clearly much slower than current German data, which shows a tenfold increase of cases every 9 days.

## Data Availability

The code is under https://github.com/matsim-org/matsim-episim, although it has been developed further since the paper was written. The code can be used together with synthetic data. The human movement data that was used for the study is unfortunately not available. The results with the synthetic data are, however, structurally similar.

http://dx.doi.org/10.14279/depositonce-9835

## 6 Acknowledgments

We thank Kai Martins-Turner for many critical and constructive comments, Dominik Ziemke for discussions, and the mathematics compute cluster of TU Berlin for CPU time.

## Footnotes

1 While we were working on the present paper, the study by (Ferguson et al. 2020) came out. Similarities and differences are discussed in the paper. Although they model at different scale (country-wide vs city-wide) and have different foci (theirs is more on the infection dynamics, ours is more on the human movement side), we believe that the results are roughly in line with each other.

2 Note that it is unclear to us if the share of critical cases is calculated as a share of the *reported* cases, or a share of the true cases. Our model simulates “true” cases.

3 This fraction is much larger than so-called “herd immunity”. We attribute this to over-shooting because of the explosive, uninhibited infection dynamics.

